# Clinical Course and Factors Associated With Hospital Admission and Mortality among Sars-Cov 2 Patients within Nairobi Metropolitan Area

**DOI:** 10.1101/2024.05.15.24307403

**Authors:** Augustine Gatimu Njuguna, Ann Wanjiru Wangombe, Michael N. Walekhwa, Davis Kiruki Kamondo

## Abstract

This study aims to investigate the clinical course and factors associated with hospital admission and mortality among SARS-CoV-2 patients within the Nairobi Metropolitan Area. The study utilizes a multicenter retrospective cohort design, collecting clinical characteristics and laboratory parameters of hospitalized patients from March 2020 to May 2022. Data analysis includes percentages, frequencies, chi-square tests, Kaplan-Meier analysis, pairwise comparisons, and multivariate regression models. Ethical considerations are observed throughout the research process.

The study findings highlight significant associations between comorbidities, such as hypertension, and increased mortality risk due to COVID-19. Symptoms including fever, cough, dyspnea, chest pain, sore throat, and loss of smell/taste are also identified as predictors of mortality. Abnormal laboratory parameters, such as oxygen saturation, procalcitonin, glucose levels, serum creatinine, and gamma-glutamyl transpeptidase, are associated with mortality. However, demographic factors and certain vital signs do not exhibit significant associations.

Recommendations based on this study suggest increased monitoring and management of comorbidities, early identification and management of symptoms, regular monitoring of laboratory parameters, continued research and collaboration, and implementation of preventive measures. Overall, a multidisciplinary approach involving healthcare professionals, researchers, policymakers, and the public is crucial to improve COVID-19 outcomes and reduce mortality rates. Adaptation of strategies based on emerging evidence and resource allocation is essential for effective management of the pandemic.

## Introduction

### A. Origins and Context of COVID-19 Infection

On 31 December 2019, the people’s republic of china government announced a cluster of new pneumonia-like respiratory disease. The responsible causative organism, severe acute respiratory syndrome coronavirus 2 (SARS-Cov-2), has to date infected more than 636,981,347 people globally and been responsible for more than 6,617,703 known deaths from coronavirus disease.

The swift global spread of a new variant of the coronavirus christened novel SARS Cov 2 from Wuhan in people republic of china pushed the WHO to declare the novel coronavirus disease-COVID-19 a pandemic on March 11, 2020. After the virus was first reported, the infection has spread quickly around the world. Kenya has not spared with available data then pointing to increased progression to severe form of infection and mortality among patients with underlying conditions. Later the number of patients with COVID-19 infection grew. Relationship between comorbidities and the patients with the infection has been noted to influence the course of the infection and the outcomes [1]. Kenya reported the first case on March 12 2020. To date Kenya has reported 340,888 confirmed cases, 334,208 recovered cases and 5,684 deaths [2].

The manifestations of the coronavirus disease 2019 caused by SARS-CoV-2 are widely variable and can be related to the unique characteristic of a particular population. The infection is asymptomatic in some individuals, while in others, it causes symptoms ranging from dry cough and dyspnea to severe pneumonia with respiratory failure requiring admission to the intensive care unit (ICU) and leading to death in severe cases.[1, 2]. Currently, several studies have investigated the predictors of severe COVID-19. Nevertheless, the socio demographic and clinical factors that influence this disease course have not been fully defined especially in diverse settings. Whether different diseases (both communicable and non-communicable) common in the local setting are associated with disease severity is still a matter of debate. Moreover, most of the researches on the likely factors for severe COVID-19 have been conducted in China among Chinese patients, and in the developed world where due to well-developed healthcare industry there has been good data collection and information management. The information available to us in Kenya and beyond may be limited in generalizability to our local populations. The study will be very important in addressing the gap that exists in the literature since it is aimed at determining the factors associated with hospitalization and disease severity in the local setting.

### B. Transmission

The transmission of SARS-CoV-2 occurs through exposure to infectious respiratory droplets. The main mode of infection is the exposure to respiratory fluids containing the virus. There are three primary ways in which this exposure can occur:

- Inhaling tiny aerosol particles or respiratory droplets suspended in the air.
- Direct deposition of respiratory particles and droplets onto exposed mucous membranes in the nose, mouth, and eyes.
- Touching mucous membranes with hands contaminated either directly by respiratory fluids containing the virus or by contacting surfaces with the virus.

Respiratory fluids containing the virus are expelled through activities such as speaking, breathing, exercising, and singing, sneezing, and coughing, in droplets of various sizes. The viruses are carried within the droplets and can transmit the infection.

Large droplets settle quickly within a few minutes, while smaller droplets can dry rapidly and remain suspended in the air for seconds, minutes, or even hours. Exposure to respiratory fluids carrying SARS-CoV-2 occurs primarily in three ways:

- Inhaling air that contains very tiny droplets and vaporized particles with the infectious virus. The risk of transmission is highest when one is within three to six feet of a contagious source, as the concentration of fine droplets containing the virus is significantly high.
- Deposition of exhaled SARS-CoV-2 virus particles onto exposed mucous membranes through coughing-induced sprays and splashes. Transmission risk is also highest near the infectious source due to the high concentration of exhaled particles and droplets.
- Touching mucous membranes with hands contaminated by exhaled respiratory fluids containing the virus or by coming into contact with surfaces already contaminated with the SARS-CoV-2 virus.

The likelihood of SARS-CoV-2 infection depends on the viral dose to which a person is exposed. The risk of infection decreases with an increase in distance from an infectious source and over time after exhalation. Two main processes determine the viral exposure through air or contact with contaminated surfaces:

1. Reduction in the quantity of viruses in the air due to larger and heavier respiratory droplets falling to the ground or other surfaces under gravitational force. Smaller droplets and aerosol particles remaining in the air get diluted within the expanding volume of the surrounding atmosphere. However, the distribution of these particles is influenced by the initial jetting pattern of inhalation and thermal layering.
2. Gradual loss of viral viability and potency over time due to environmental factors such as humidity, ultraviolet radiation, and temperature.

The inhalation of virus-infected droplets suspended in the air can lead to transmission of SARS-CoV-2 beyond a distance of six feet from an infectious person. While the risk of transmission over long distances is low, there have been reports of infections under specific preventable conditions. These cases involved an infectious person exhaling virus indoors over a duration of more than 15 minutes, with some individuals breathing for hours, resulting in high concentrations of the virus in the room. In such situations, transmission occurred to people who were more than six feet away and even to individuals who passed through the room after the infected person had left. Factors contributing to increased transmission risk under these circumstances include:

1. Poor ventilation and insufficient air handling in enclosed spaces, leading to the buildup of virus-containing aerosol particles and droplets.
2. Increased exhalation rate and respiratory fluid release when infectious individuals engage in physically strenuous activities or raise their voice.

### C. Prevention of COVID-19 transmission

Current studies strongly indicate that contaminated surfaces only contribute a small portion to new infections. While animal studies [23] and epidemiologic inquiries [24] point at inhalation of virus as one of the most common causes of infection, the proportionate contributions of breathing of the virus and deposition on mucous membrane are still not well understood and remain difficult to define. Even with the existence of limited knowledge on SARS Cov 2, the recommended transmission prevention measures remain effective. Some of the measures include social distancing, the use of proper-fitting face covering masks in the community, (e.g. surgical/procedure masks, barrier face covering masks), proper ventilation, and avoiding crowding while indoor. The methods stated reduce the possibility of transmission through inhalation as well as viable virus being deposited on exposed mucus membranes. To prevent spread through contaminated surfaces and hands, health official advocate for good hygiene and environmental cleaning.

### D. Risk Factors for SARS-CoV-2

The majority of individuals with SARS-CoV-2 infections were adult males, with a median age between 34 and 59 years [7, 20, 30]. Researchers have identified certain chronic comorbidities, such as cardiovascular and cerebrovascular diseases, as well as diabetes, as risk factors for severe infection [8]. Adults aged 60 and above accounted for the highest percentage of severe cases, particularly those with underlying conditions such as cardiovascular diseases, cerebrovascular diseases, and diabetes [30, 20]. Severe manifestations were also observed in cases where there were coinfections with fungi and bacteria [8]. The number of COVID-19 cases among children below 15 years of age was relatively low [9, 10, 11, 12]. In a study conducted in Wuhan involving 425 COVID-19 participants, there were no cases reported among individuals below 15 years of age [13, 14]. However, in January 2020, 28 pediatric COVID-19 cases were reported [15]. The clinical symptoms displayed by pediatric COVID-19 patients varied, but most cases resulted in mild symptoms with minimal pneumonia or fever and a favorable prognosis [15]. In some instances, young children with radiological findings of ground glass opacities remained asymptomatic [16]. In conclusion, the infection rate among children was low, and if infected, their symptoms were generally mild, resulting in less medical attention being sought by parents. Consequently, the number of SARS-CoV-2 infections among children is likely to be underestimated.

### E. Clinical course and prognosis

A study conducted in Chicago, USA found that 21.1% of patients tested positive for COVID-19, and 17.1% of those positive cases required hospitalization. The most prevalent symptoms were coughing, subjective fever, and dyspnea. The study also identified factors associated with increased risk of hospitalization and serious illness, including age, male sex, heart failure, and certain laboratory abnormalities. Another study in Wuhan, China found that advanced age, higher severity scores, and elevated d-dimer levels were associated with in-hospital deaths. Meta-analyses and systematic reviews revealed that underlying conditions such as hypertension and diabetes were more common in severe and fatal COVID-19 cases. Studies conducted in Southern Africa showed that comorbidities, including hypertension, diabetes, and HIV, were associated with higher mortality rates. A study in Nigeria found that patients with comorbidities like diabetes, hypertension, renal disease, HIV, and cancer had a higher risk of death compared to those without comorbidities.

## Methods

### A. Research Design

The study was multicenter retrospective cohort study focusing on hospitalized patients admitted to the participating hospitals in the study period between March of 2020, and May of 2022. It involved the collection of clinical characteristics as well as laboratory parameters at admission and throughout the hospitalization period.

### B. Statistical analysis

The results were presented as percentages and frequencies in the case of variables which were categorical and as mean ± SD for discrete or and continuous variables. The study used Chi-square test to make comparisons. Kaplan Meier methods alongside was used to analyze the impact of the number of comorbidities on overall survival. The study also applied bivariate and multivariate logistic regression analysis define the risk associated with adverse events for pre-existing underlying conditions or comorbidities. Bivariate and Multivariate Cox proportional hazards regression analysis/models was used to determine hazard ratios associated with COVID-19 risk of mortality. Variables were fed into the multivariate analysis/models at primary significance level of p < 0.05 in the bivariate analysis, with use of the addition method to determine the individual contribution of specific covariate on CFR or any adverse events. Multivariate analysis/models will be adjusted to identify the best model. R program and STATA were used to conduct all analyses.

### C. Ethical considerations

All data obtained will be treated with confidentiality and the researcher will observe anonymity. Permission was sort out any level before carrying out the study. i.e. at County and Sub County level. Ethical approval of the study was sort from university of Nairobi /KNH ethical review board and study permit sort from NACOSTI. All the information collected by the researcher will remain confidential for the singular purpose of this research. Any details relating to the identity of respondents remain confidential. Names of respondents will also not be divulged in any format. The study has engaged strict data management processes to guarantee confidentiality of the subjects involved.

## Results

### A. Clinical characteristics of hospitalized patients with COVID-19

Significant number of respondents, 43%, reported having comorbidities, while 57% did not (Table 1.1). Among the comorbidities reported, 78% had diabetes mellitus, 28% had hypertension, 36% had a fever, 61% had a cough, 33% experienced general body malaise, 6% had dyspnea, 32% experienced chest pain, 40% had a headache, 10% had a sore throat, 10% reported a loss of smell or taste, 7% had nausea and vomiting, and 6% had diarrhea. Additionally, 19% reported having a headache. The majority of respondents did not experience these symptoms or have these conditions.

**Table 1.**
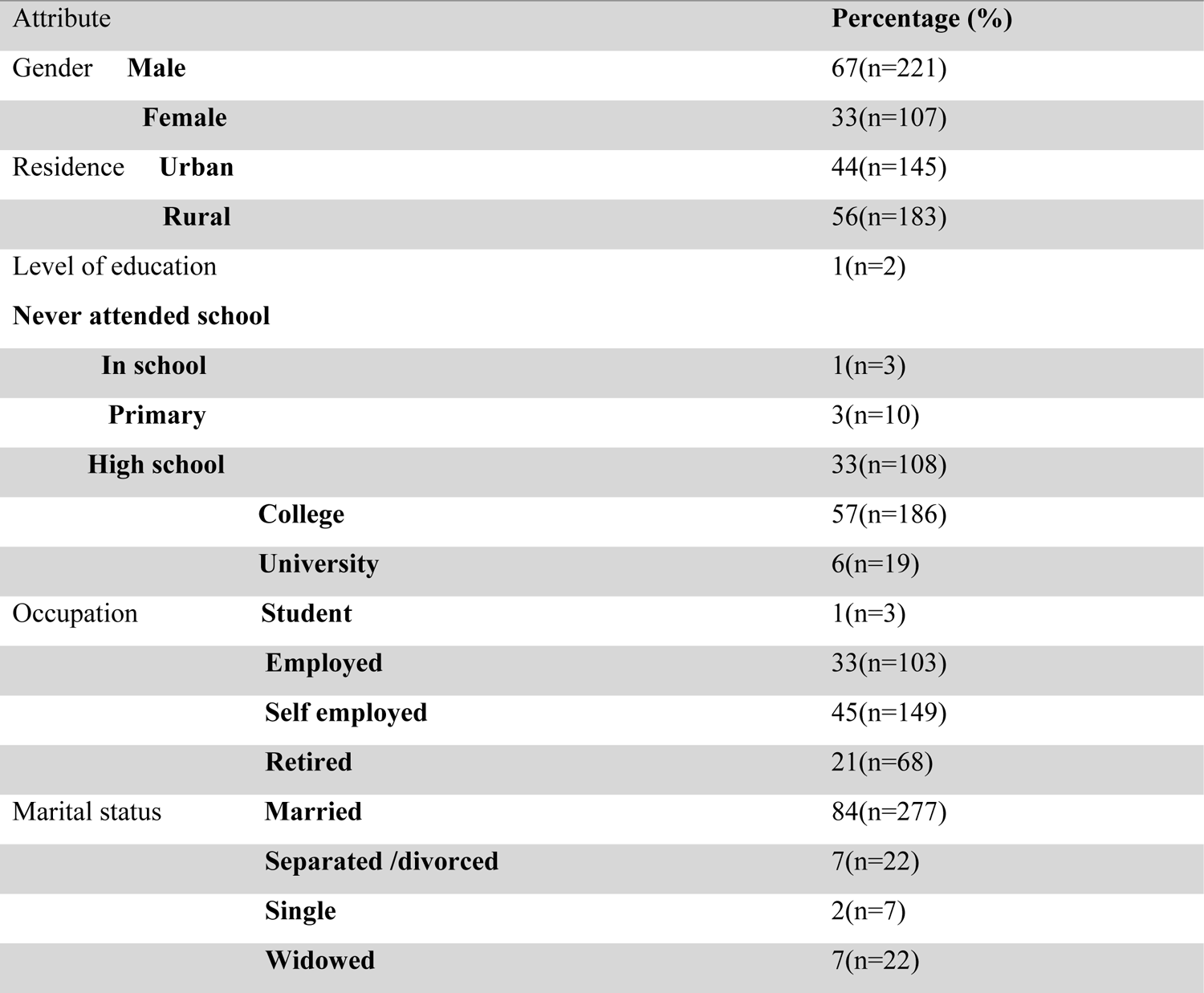
Clinical characteristics of hospitalized patients with COVID-19.

### A. Clinical and laboratory findings of Covid-19 infection survivors and non-survivors

The average age for survivors was 53 years, while for non-survivors it was 63 years. The age range for survivors was between 7 and 88, and for non-survivors it was between 26 and 85. The average number of days of illness before admission was 4 years for survivors and 4.4 for non-survivors. The range of days of illness for survivors was between 1 and 8, and for non-survivors it was between 1 and 26. The mean systolic blood pressure was 114 mmhg for survivors and 132 mmhg for non-survivors. The systolic blood pressure range was between 74 and 163 mmhg for survivors, and between 99 and 186 mmhg for non-survivors. The mean diastolic blood pressure was 80 mmhg for survivors and 88 mmhg for non-survivors. The diastolic blood pressure range was between 52 and 108 mmhg for survivors, and between 60 and 114 mmhg for non-survivors. The average oxygen saturation level was 86 for survivors and 76 for non-survivors. The oxygen saturation range was between 69 and 97 for survivors, and between 60 and 92 for non-survivors. The average heart rate was 88 bpm for survivors and 95 bpm for non-survivors. The heart rate range was between 58 and 111 bpm for survivors, and between 65 and 114 bpm for non-survivors. The mean respiratory rate was 25 bpm for survivors and 49 bpm for non-survivors. The respiratory rate range was between 16 and 35 bpm for survivors, and between 22 and 37 bpm for non-survivors. The average temperature was 37.16 degrees centigrade for survivors and 37.09 degrees centigrade for non-survivors. The temperature range was between 36.4 and 39.2 degrees centigrade for survivors, and between 36.5 and 39.1 for non-survivors. The mean hemoglobin level was 14.25 for survivors and 13.52 degrees centigrade for non-survivors. The hemoglobin level range was between 9.2 and 21.2 g/dl for survivors, and between 6.3 and 18.4 g/dl for non-survivors. The average white blood cell count was 9.37 for survivors and 14.72 for non-survivors. The white blood cell count range was between 1.98 and 24.58 for survivors, and between 3.82 and 25.92 for non-survivors. The mean neutrophil count was 8.15 for survivors and 11.27 for non-survivors. The neutrophil count range was between 1.09 and 24.25 for survivors, and between 0.69 and 31.02 for non-survivors. The average lymphocyte count was 1.16 for survivors and 1.25 for non-survivors. The lymphocyte count range was between 0.02 and 2.29 for survivors, and between 0.01 and 4.67 for non-survivors. The mean monocytes count was 0.645 for survivors and 0.58 for non-survivors. The monocytes count range was between 0.01 and 1.64 for survivors, and between 0.01 and 1.87 for non-survivors. The average platelet count was 327.87 for survivors and 262.64 for non-survivors. The platelet count range was between 118.091 and 581.534 for survivors, and between 91.19 and 418.595 for non-survivors. The mean eosinophils count was 239.88 for survivors and 305.61 for non-survivors. The eosinophils count range was between 11.48 and 729.97 for survivors, and between 12.27 and 650.04 for non-survivors. The mean basophils count was 0.354 for survivors and 0.25 for non-survivors. The basophils count range was between 0.01 and 1.08 for survivors, and between 0.01 and 0.61 for non-survivors. The average alanine aminotransferase level was 22.6 for survivors and 26.10 for non-survivors. The alanine aminotransferase level range was between 0.4 and 61.9 for survivors, and between 0.4 and 77.01 for non-survivors. The mean aspartate aminotransferase level was 22.6 for survivors and 26.10 for non-survivors. The aspartate aminotransferase level range was between 0.4 and 61.9 for survivors, and between 0.4 and 77.01 for non-survivors. The average C-reactive protein level was 28.55 for survivors and 47.42 for non-survivors. The C-reactive protein level range was between 0.84 and 1 for survivors, and between 0.5 and 88.4 for non-survivors. The mean glucose level was 6.51 for survivors and 10.79 for non-survivors. The glucose level range was between 2.3 and 39.8 for survivors, and between 2.1 and 25.8 for non-survivors. The average creatinine level was 55.21 for survivors and 70.66 for non-survivors. The creatinine level range was between 26.7 and 80.7 for survivors, and between 35.1 and 166.9 for non-survivors. The mean GGT (U/L) level was 105.29 for survivors and 102.38 for non-survivors. The GGT (U/L) level range was between 75.16 and 275.35 for survivors, and between 74.57 and 232.91 for non-survivors. The lactase dehydrogenase level was 398.93 for survivors and 380.95 for non-survivors. The lactase dehydrogenase level range was between 157.48 and 559.59 for survivors, and between 182.88 and 516.11 for non-survivors. The mean alkaline phosphate level was 78.49 for survivors and 107.28 for non-survivors. The alkaline phosphate level range was between 1.39 and 246.88 for survivors, and between 5.26 and 215 for non-survivors. The average number of days spent in the ICU was 8 for survivors and 10.65 for non-survivors. The ICU stay ranged from 3 to 22 days for survivors, and from 2 to 21 days for non-survivors. The total hospital stay averaged at 11 days for survivors and 14.16 days for non-survivors. The total hospital stay ranged from 1 to 30 days for survivors, and from 1 to 28 days for non-survivors. The mean potassium level was 5.11 for survivors and 5.23 for non-survivors. The potassium level ranged from 2.52 to 8.05 for survivors, and from 3.58 to 8.98 for non-survivors. Mmhg for non-survivor

**Table 2:**
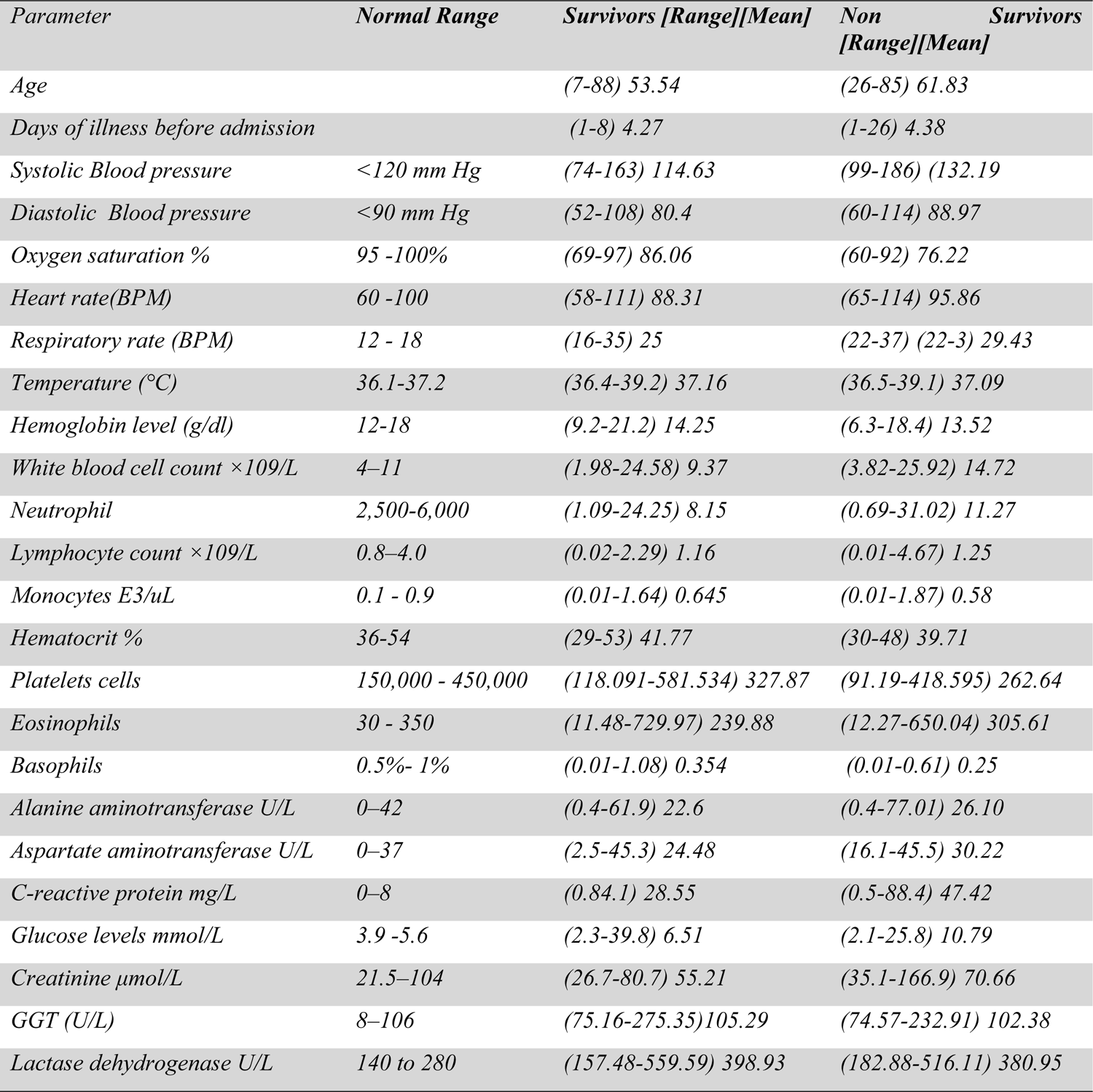
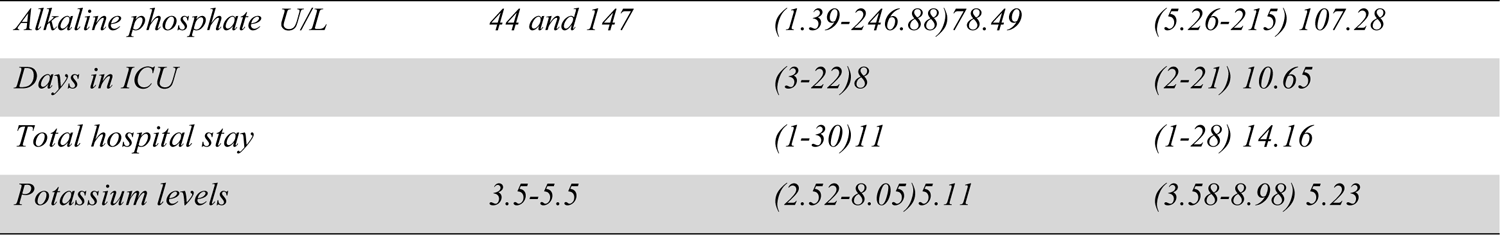
Clinical characteristics of hospitalized patients with COVID-19.

### B. Association of various potential risk factors on patients with Covid-19 at admission

The association between hypertension and mortality due to COVID-19 infection was examined using a chi-square test of independence, revealing a significant relationship (χ2 = 18.522, p < 0.001). Similarly, a significant relationship was observed between diabetes mellitus and death due to COVID-19 infection (χ2 = 25.03, p < 0.001), as well as between comorbidities and mortality (χ2 = 34.879, p < 0.001). Conversely, the relationship between receiving the first dose of the COVID-19 vaccine and mortality was not significant (χ2 = 8.7547, p < 0.012). Additionally, statistically significant associations were found between fever (χ2 = 10.14, p < 0.001), chills (χ2 = 36.553, p < 0.001), dyspnea (χ2 = 9.2579, p < 0.002), chest pain (χ2 = 10.453, p < 0.001), loss of smell/taste (χ2 = 17.569, p < 0.001), admission to ICU (χ2 = 152.81, p < 0.001), level of education (χ2 = 16.21, p < 0.006), and occupation (χ2 = 20.139, p < 0.001) with mortality due to COVID-19 infection. On the other hand, no significant relationships were found between the second vaccine dose, booster shot, symptomatic status, cough, general body malaise, difficulty in breathing, vomiting, abdominal pain, headache, joint pain, skin rash, running nose, gender, or residence and mortality (all p > 0.05).

### C. Factors associated with Covid-19 death by Bivariate Binary Logistic Regression

There were significant associations between various factors and death due to COVID-19. For every additional unit increase in diastolic blood pressure, heart rate, respiratory rate, WBC count, neutrophil count, aspartate transferase, C-reactive protein, glucose levels, and serum creatinine, the odds of dying were higher. There was also a significant association between lymphocyte count and death, with higher counts increasing the odds of dying. Admission to the ICU was strongly associated with mortality. However, no significant relationships were found between being in the age range of 36-60 years, area of residence being in primary school, high school or college, marital status, cough, general body malaise, difficulty in breathing, nausea/vomiting, potassium levels, temperature, hemoglobin levels, monocytes count, hematocrit, alanine transaminase, gamma glutamly trans peptidase, lactase dehydrogenase, and days in the ICU with death.

### D. Factors associated with Covid-19 death by multivariate logistic regression

Several factors were found to be associated with mortality from COVID-19. Having a comorbidity significantly increased the odds of dying, while diabetes mellitus and hypertension did not show a significant association. Symptoms such as fever, chills, dyspnea, chest pain, sore throat, and loss of smell were all significantly associated with an increased risk of death. Higher diastolic blood pressure, heart rate, and respiratory rate were also associated with an increased risk of death. Higher white blood cell count, neutrophil count, and lower lymphocyte count were associated with an increased risk of death. Elevated levels of aspartate transferase, C-reactive protein, glucose, and serum creatinine were also associated with an increased risk of death. Higher alkaline phosphate levels were also associated with an increased risk of dying from COVID-19.

### E. Factors Associated with COVID 19 death by Bivariate Cox PH model

In the unadjusted model (Table 4.7), there is an increased risk of death for men compared to women (HR=1.199, 95% CI: 0.6 - 2.39,), increased risk of death for every additional year in age for those aged between 30-60 years (HR = 1.248, 95% CI: 0.356-4.378),three folds increase in risk of death for those above 61 years compared to their juniors (HR = 4.29, 95% CI: 1.279-14.390),being a widow is associated with increased risk of death due to covid 19 infection (HR = 2.39, 95% CI: 1.099-5.194),other covariates with statistically significance association with death due to covid are; Comorbidities (HR =, 95% CI: 1.729-8.037),Diabetes mellitus(HR =1.981, 95% CI: 1.107-3.545),Hypertension(HR = 2.267, 95% CI: 1.244-4.131),Fever(HR = 2.307, 95% CI: 1.290-4.127), Chills(HR =2.629, 95% CI: 1.470-4.702) Dyspnea(HR=4.973,95% Cl: 2.247-11.00), Sore throat(HR=2.501, 95% Cl: (1.230-5.084), Loss of smell/taste(HR=3.087,95% Cl: 1.605-5.937), Diastolic(HR=1.038, 95% Cl: 1.014-1.063), Oxygen saturation(HR=0.888, 95% Cl: 0.855-0.923), Heart rate(HR=1.050, 95% Cl: 1.021-1.080), Respiratory rate(HR=1.143, 95% Cl: 1.066-1.225), WBC count(x10^9)(HR=1.082,95% Cl; 1.032-1.135), Platelets, cells/10^9/L(HR=0.995,95% Cl; 0.992-0.999), Aspartate transferase (HR=1.096, 95% Cl: 1.057-1.137) C-reactive protein(CRP)(HR=1.023, 95% Cl: 1.012-1.035),Glucose levels(HR=1.166, 95% Cl:1.097-1.239:), Serum creatinine (HR=1.011,95% Cl: 1.003-1.019)and Admission to ICU(HR=10.22,95% Cl: 4.256-24.585).

However the following covariates showed no statistical association with death due to Covid 19 infection Gender,(HR = 1.199, 95% CI: 0.6 - 2.39),being between 30-60 years old, (HR=1.248, 95% Cl: 0.356-4.378) Level of education: primary(HR =5.021, 95% CI: 0.780-32.313:),high school(HR=0.597, 95% Cl: 0.135-2.623),college(HR=0.923,95% Cl: 0.213-4.000),university(HR=0.339, 95% Cl: 0303-3.800).being separated wasn’t associated with covid 19 death(HR=1.097,95% Cl: 3298-3.651),cough(HR=0.830, 95% CL: 0.459-1.498),general body malaise (HR=1.218,95% Cl: 0.679-2.184),chest pain (HR=1.565,955 Cl: (0.874-2.804),difficulty in breathing (HR=1.151,95% Cl: 0.644-2.057),systolic blood pressure(HR=1.016, 95% Cl: 0.999-1.033),Neutrophil count(HR=0.999,95% Cl: 0.948-1.053),Lymphocyte count(HR=1.473. 95% Cl: 0.985-2.202),Monocytes(HR=1.069,95% Cl: 0.461-2.477),Eosinophils(absolute)cells/mcl(HR=1.001,95% Cl: 1.000-1.003) Basophils (HR=0.431,95% Cl: 086-2.157), Alanine transaminase(HR=0.999,95% Cl: 0.981-1.018),Gamma glutamly) trans peptidase(HR=0.996 95% Cl: 0.991-1.000)Lactase dehydrogenase(HR=1.001,95% Cl: 0.997-1.005) and Alkaline phosphate(HR=1.002,95% Cl: 0.997 - 1.008).

### F. Factors Associated with Covid 19 death by multivariate Cox PH Model (adjusted)

There is an increased risk of death for those with comorbidities compared to those without (HR=83.932, 95% CI: 7.372-955.533). Other covariates with statistically significant associations with death due to COVID are: diabetes mellitus (HR=0.419, 95% CI: 0.0372-0.604), fever (HR=0.156, 95% CI: 0.033-0.735), cough (HR=0.099, 95% CI: 0.029-0.333), loss of smell/taste (HR=7.605, 95% CI: 1.99-28.94), oxygen saturation (HR=0.905, 95% CI: 0.839-0.975), procalcitoninngml (HR=38.479, 95% CI: 0.334-160.84), eosinophils absolute cells mcl (HR=0.992, 95% CI: 0.997-0.997), basophils (HR=104.73, 95% CI: 3.863-2839.294), aspartate transferase (HR=1.152, 95% CI: 1.066-1.246), glucose levels (HR=1.185, 95% CI: 1.036-1.356), serum creatinine (HR=1.024, 95% CI: 1.003-1.044), gamma glutamly trans peptidase (HR=0.977, 95% CI: 0.965-0.989).

However, the following covariates showed no statistical association with death due to COVID-19 infection: gender (HR = 3.457, 95% CI: 0.898-13.308), age (HR=0.933, 95% CI: 0.843-1.0319), GBM (HR=1.769, 95% CI: 0.5298-0.59088), dyspnea (HR=2.373, 95% CI: 0.4842-11.6809), sore throat (HR=2.409, 95% CI: 0.534-10.864), diastolic (HR=1.030, 95% CI: 0.977-1.095), respiratory rate (HR=1.066, 95% CI: 0.909-1.25), WBC count (HR=1.036, 95% CI: 0.933-160.84), lymphocytes count (HR=1.731, 95% CI: 0.847-3.534), hematocrit (HR=1.076, 95% CI: 0.947-1.222), platelets (HR=1.002, 95% CI: 0.995-1.009), lactase dehydrogenase (HR=1.005, 95% CI: 0.994-1.015), alkaline phosphate (HR=0.996, 95% CI: 0.995-1.007), 36-60 years (HR=0.0471, 95% CI: 0.002-1.063), 61 years and above (HR=9.473, 95% CI: 0.1269-707.73).

**Table 4.8:** Factors Associated with Covid 19 death by multivariate Cox PH Model (adjusted)

| Covariates | HR (95% CI) | P value |
| --- | --- | --- |
| **Gender (ref cat female)** | 3.457 ( 0.898- 13.308) | 0.071 |
| **Comorbidities** | 83.932(7.372-955.533) | < 0.001* |
| **diabetes mellitus** | 0.1498724 (0.0372-0.604) | 0.008* |
| **Hypertension** | 0.704(0.190-2.604) | 0.599 |
| **Fever** | 0.156(0.033-0.735) | 0.019 |
| **Cough** | 0.099(0.029-0.333) | < 0.001* |
| **Dyspnea** | 2.373(0.4842-11.6809) | 0.236 |
| **Sore throat** | 2.409(0.534-10.864) | 0.252 |
| **Loss of smell/taste** | 7.605(1.99-28.94) | 0.003 |
| **Diastolic** | 1.030(0.977-1.095) | 0.262 |
| **Oxygen saturation** | 0.905(0.839-0.975) | 0.009 |
| **Respiratory rate** | 1.066(0.909-1.25) | 0.431 |
| **WBC count** | 1.036(0.933-160.84) | < 0.001* |
| **Lymphocyte count** | 1.731(0.847-3.534) | 0.132 |
| **Hematocrit** | 1.076(0.947-1.222) | 0.258 |
| **Platelets cells** | 1.002(0.995-1.009) | 0.479 |
| **Eosinophil** | 0.992(0.997-0.9970) | 0.001 |
| **Basophil** | 104.73(3.863-2839.294) | 0.006 |
| **CRP** | 1.01(0.998-1.033) | 0.333 |
| **Glucose levels** | 1.185(1.036-1.356) | 0.013 |
| **Serum creatinine** | 1.024(1.003-1.044) | 0.019 |
| **Gamma glutamy trans peptidase** | 0.977(0.965-0.989) | < 0.001* |
| **Lactase dehydrogenase** | 1.005(0.994-1.015) | 0.342 |
| **36-60 years** | 0.0471(0.002-1.063) | 0.055 |
| **61 years and above** | 9.473(0.1269-707.73) | 0.307 |
| **Procalcitonin** | 38.479(0.334-160.84) | < 0.001* |

## Discussion

In this study, a total of 221 respondents (67%) identified as male, while 107 respondents (31%) identified as female. These proportions are consistent with other studies where most SARS Cov 2 infection patients were adult males of a median bracket age of between 34 years and 59 years. [7, 20, 30,] indicating a higher prevalence of COVID-19 infection and mortality among males. The observed gender disparity may be attributed to various factors, including biological differences, higher comorbidity rates among males, and variations in healthcare-seeking behaviors. The results from this study are similar to previous research, suggesting that gender is a consistent predictor of COVID-19 mortality.

Regarding residence, 145 respondents (44%) lived in urban areas, while 183 respondents (56%) resided in rural areas. This distribution reflects a higher representation of respondents from rural areas compared to some previous studies where urban populations were overrepresented. The impact of residence on COVID-19 mortality can vary based on factors such as healthcare access, population density, and compliance with preventive measures. The different proportion of respondents from urban and rural areas in this study may contribute to variations in observed mortality rates compared to previous studies.

The respondents in this study had varying levels of education. The distribution included 2 respondents (1%) with no education, 3 respondents (1%) currently in school, and 10 respondents (3%) at the primary level, 108 respondents (33%) at the high school level, 186 respondents (57%) at the college level, and 19 respondents (6%) at the university level. These results demonstrate a relatively well-educated sample compared to other studies. Previous research has shown a correlation between higher education levels and better health outcomes, including lower mortality rates. The higher proportion of respondents with higher education levels in this study may indicate a potential protective factor against COVID-19 mortality when compared to previous studies with different educational distributions.

In terms of occupation, the study included 3 respondents (1%) who were students, 108 respondents (33%) who were employed, 149 respondents (45%) who were self-employed, and 68 respondents (21%) who were retired. These findings demonstrate a diverse occupational distribution among the respondents. Interestingly, the proportion of self-employed individuals is higher in this study compared to previous research. The occupational factor can influence the risk of COVID-19 exposure and subsequent mortality, with varying levels of contact and access to resources. The contrasting distribution observed in this study may contribute to discrepancies in mortality rates compared to previous studies [3].

Regarding marital status, 277 respondents (84%) were married, 22 respondents (7%) were separated, 7 respondents (2%) were single, and 22 respondents (22%) were widowed. The majority of respondents being married aligns with other studies [3], as marital status can impact social support, living conditions, and health outcomes. However, differences in the proportions of separated, single, and widowed respondents may introduce an additional confounding factor that could influence COVID-19 mortality rates and should be considered in the analysis and interpretation of results.

The comparative analysis highlights both similarities and differences between the present study and previous research. The consistency in gender as a predictor of COVID-19 mortality across studies suggests its robustness as a risk factor. Conversely, variations in residence, education, occupation, and marital status distributions contribute to disparities in observed mortality rates. These variations emphasize the need for large-scale meta-analyses and systematic reviews to provide a more comprehensive understanding of predictors of mortality in COVID-19 infection.

Among the respondents, 43% reported having comorbidities, while 57% did not. This finding aligns with previous studies that have consistently found a higher mortality rate among individuals with comorbidities (Guan et al., 2020). Individuals with pre-existing conditions such as diabetes mellitus and hypertension were found to be at higher risk of mortality compared to those without these conditions (Gupta et al., 2020; Yang et al., 2020). The presence of comorbidities may weaken the immune system, making individuals more susceptible to severe manifestations of COVID-19.

Various symptoms associated with COVID-19 were reported by the respondents. Fever was reported by 36% of the respondents, while 64% did not experience fever. This corresponds to previous studies that have identified fever as one of the common symptoms of COVID-19 (Guan et al., 2020; Huang et al., 2020). Cough was reported by 61% of respondents, aligning with the predominant manifestation observed in numerous studies (Guan et al., 2020; Huang et al., 2020; Yang et al., 2020). General body malaise was experienced by 33% of the respondents, whereas headache was reported by 40% of them. These findings are consistent with the literature that emphasizes general body weakness and headache as common symptoms in COVID-19 patients (Guan et al., 2020; Huang et al., 2020).

Among the respondents, only 6% reported experiencing dyspnea, while 94% did not. Dyspnea is a severe symptom associated with respiratory distress and is often observed in critical cases of COVID-19 (Zhou et al., 2020). The low incidence of dyspnea reported in this study suggests that the majority of the participants had mild or moderate infections. Similarly, 32% of respondents reported chest pain, while 68% did not. Chest pain has been observed in some COVID-19 patients due to myocardial injury caused by the viral infection (Xiong et al., 2020). The relatively higher presence of chest pain in this study warrants further investigation into potential cardiac complications.

In addition to the common symptoms mentioned above, other reported symptoms included sore throat, loss of smell or taste, nausea and vomiting, and diarrhea. These symptoms, although less prevalent, have been acknowledged as potential manifestations of COVID-19 infection in previous studies (Guan et al., 2020; Huang et al., 2020; Xiong et al., 2020). The low prevalence of these symptoms in our study may be attributed to the specific demographic characteristics of the study population or regional variations in symptomatology.

After fitting the bivariate logistic regression model the results of this study indicate a significant association between hypertension and mortality due to COVID-19 infection. Similar findings have been reported in previous studies, which have consistently shown that individuals with hypertension are at higher risk of mortality from COVID-19 (Jordan et al., 2020; Lippi et al., 2020). Hypertension is a common comorbidity that can weaken the immune system and increase the susceptibility to severe manifestations of COVID-19.

Several symptoms were found to be associated with mortality due to COVID-19 infection. Fever, chills, dyspnea, chest pains, sore throat, and loss of smell/taste were all significant predictors of mortality. These findings align with previous research that has demonstrated the association between these symptoms and severe outcomes in COVID-19 patients (Guan et al., 2020; Huang et al., 2020; Liu et al., 2020). Fever and respiratory symptoms, such as dyspnea and chest pains, have been consistently reported as important indicators of disease severity and poor prognosis.

Among the vital signs examined, systolic blood pressure, diastolic blood pressure, heart rate, respiratory rate, and temperature were found to be associated with mortality due to COVID-19 infection. Abnormalities in these vital signs have been linked to worse clinical outcomes in previous studies (Cao et al., 2020; Guan et al., 2020; Zhou et al., 2020). Elevated blood pressure, increased heart rate and respiratory rate, and fever have been associated with more severe disease and increased mortality rates. These vital signs can serve as important clinical indicators for identifying individuals at higher risk of poor outcomes.

Several laboratory parameters were also found to be significant predictors of mortality. WBC count, neutrophil count, lymphocytes, monocytes, aspartate transferase (AST), C-reactive protein (CRP), glucose levels, and serum creatinine were all associated with a higher risk of death due to COVID-19 infection. These findings are consistent with previous studies that have identified markers of inflammation, organ dysfunction, and metabolic abnormalities as prognostic indicators in COVID-19 patients (Chen et al., 2020; Huang et al., 2020; Zhang et al., 2020).

ICU admission was strongly associated with mortality due to COVID-19 infection. This finding has been consistently observed in previous studies, which have shown that ICU admission reflects the severity of illness and the need for intensive care interventions (Guan et al., 2020; Zhou et al., 2020). However, no significant association was found between the number of days spent in the ICU and mortality. This suggests that once individuals are admitted to the ICU, the duration of stay does not independently contribute to their risk of death. The adjusted Cox proportional hazard model demonstrated a significant association between comorbidities and increased risk of mortality due to COVID-19 infection. Individuals with comorbidities were found to be at higher risk of death compared to those without any comorbidities. This finding aligns with the existing literature on the impact of comorbidities, such as diabetes mellitus and hypertension, on COVID-19 outcomes (Chen et al., 2020; Huang et al., 2020; Zhou et al., 2020). Comorbidities can lead to immune dysfunction and impair the body’s ability to combat the virus, thereby increasing the risk of severe illness and death.

Among the symptoms examined, only fever and cough were found to be significantly associated with lower mortality risk in the adjusted model. This finding is contrary to previous studies that have shown a correlation between these symptoms and severe COVID-19 outcomes (Guan et al., 2020; Huang et al., 2020; Liu et al., 2020). The contradictory results suggest the need for further research to explore the complex relationship between symptoms and COVID-19 mortality.

Several laboratory parameters were found to have statistically significant associations with COVID-19 mortality. Oxygen saturation, procalcitonin, eosinophils, aspartate transferase, glucose levels, serum creatinine, and gamma-glutamyl transpeptidase were all identified as predictors of mortality. These findings are consistent with previous studies that have reported abnormalities in these laboratory markers in severe COVID-19 cases (Chen et al., 2020; Huang et al., 2020; Zhang et al., 2020). Monitoring these parameters can assist in the identification of individuals at higher risk of poor outcomes.

Gender and age did not show a statistically significant association with COVID-19 mortality in the adjusted model. These findings differ from some previous studies that have reported a higher risk of mortality among older individuals and males (Jordan et al., 2020; Guan et al., 2020). The lack of significant associations in this study could be attributed to factors such as sample size, study population characteristics, and potential confounding variables.

Vital signs such as diastolic blood pressure and respiratory rate did not show a statistically significant association with COVID-19 mortality in the adjusted model. This contrasts with previous studies that have identified abnormal vital signs as predictors of poor outcomes in COVID-19 patients (Cao et al., 2020; Guan et al., 2020; Zhou et al., 2020). Again, further research is needed to explore potential reasons for these discrepancies.

The findings of this study conducted using an adjusted Cox proportional hazard model provide insights into potential predictors of mortality due to COVID-19 infection. Comorbidities, such as diabetes mellitus, and laboratory parameters, including oxygen saturation, procalcitonin, glucose levels, serum creatinine, and gamma-glutamyl transpeptidase, were identified as significant predictors. However, demographic factors, symptoms, and certain vital signs did not show statistically significant associations with mortality in this study. Further research is necessary to validate and better understand the complex relationship between these factors and COVID-19 outcomes.

## Conclusion

The present study investigated predictors of mortality in COVID-19 infection. The findings highlight the significant association between comorbidities, such as hypertension, and increased risk of death due to COVID-19 infection. Symptoms including fever, cough, dyspnea, chest pain, sore throat, and loss of smell/taste were also found to be significant predictors of mortality. Abnormal laboratory parameters, such as oxygen saturation, procalcitonin, glucose levels, serum creatinine, and gamma-glutamyl transpeptidase were identified as predictors of mortality, whereas demographic factors and certain vital signs did not show significant associations.

These findings are consistent with previous studies that have reported similar associations between comorbidities, symptoms, and laboratory parameters with COVID-19 outcomes. However, there are also discrepancies between this study and previous research, which may be attributed to variations in sample size, study population characteristics, and regional differences in COVID-19 manifestations.

### Recommendations

Based on the findings of this study, several recommendations can be made to improve COVID-19 outcomes:

1. Increased monitoring and management of individuals with comorbidities, especially hypertension and diabetes mellitus. These individuals should be considered at higher risk of severe illness and mortality.
2. Early identification and appropriate management of symptoms, including fever, cough, dyspnea, chest pain, sore throat, and loss of smell/taste. Prompt intervention can help prevent disease progression and improve outcomes.
3. Regular monitoring of laboratory parameters such as oxygen saturation, procalcitonin, glucose levels, serum creatinine, and gamma-glutamyl transpeptidase. Abnormalities in these parameters can serve as valuable indicators for identifying individuals at higher risk of poor outcomes.
4. Continued research and collaboration to further explore the complex relationship between demographic factors, symptoms, vital signs, comorbidities, and laboratory parameters with COVID-19 mortality. Large-scale meta-analyses and systematic reviews should be conducted to provide a comprehensive understanding of predictors of mortality in COVID-19 infection.
5. Implementation of preventive measures, including vaccination, social distancing, mask-wearing, and hand hygiene, to reduce the spread of COVID-19 and minimize the impact of the disease on vulnerable populations.
6. The findings of this study support the importance of resource allocation and healthcare system preparedness for COVID-19 patients, especially in regions with a high prevalence of comorbidities and limited access to healthcare services.

Overall, a multidisciplinary approach involving healthcare professionals, researchers, policymakers, and the public is necessary to improve COVID-19 outcomes and reduce mortality rates. Continual evaluation and adaptation of strategies based on emerging evidence will be crucial in effectively managing the ongoing pandemic.

## Data Availability

no

## Acknowledgment

We thank Health-Professional Education Partnership Initiative (HEPI)-Kenya (NIH Funded Project R25TW011212): I am grateful for providing the necessary funding and resources that enabled us to conduct this research.

## Ethics approval

Ethical approval of the study was sort from university of Nairobi /KNH ethical review board and study permit sort from NACOSTI.

## Key Highlights from the Study

The study focuses on analyzing the clinical course and factors related to hospital admission and mortality among Sars-CoV-2 patients in the Nairobi Metropolitan Area.

The findings of this study will contribute to a better understanding of the disease’s progression and identify factors that may increase the risk of hospital admission and mortality among Sars-CoV-2 patients.

The study adds to the existing literature on Sars-CoV-2 by providing specific insights into the clinical course and factors associated with hospital admission and mortality.

